# A chest radiography-based artificial intelligence deep-learning model to predict severe Covid-19 patient outcomes: the CAPE (Covid-19 AI Predictive Engine) Model

**DOI:** 10.1101/2020.05.25.20113084

**Authors:** Charlene Liew, Jessica Quah, Han Leong Goh, Narayan Venkataraman

## Abstract

**Background:** Chest radiography may be used together with deep-learning models to prognosticate COVID-19 patient outcomes

**Purpose:** T o evaluate the performance of a deep-learning model for the prediction of severe patient outcomes from COVID-19 pneumonia on chest radiographs.

**Methods:** A deep-learning model (CAPE: Covid-19 AI Predictive Engine) was trained on 2337 CXR images including 2103 used only for validation while training. The prospective test set consisted of CXR images (n=70) obtained from RT-PCR confirmed COVID-19 pneumonia patients between 1 January and 30 April 2020 in a single center. The radiographs were analyzed by the AI model. Model performance was obtained by receiver operating characteristic curve analysis.

**Results:** In the prospective test set, the mean age of the patients was 46 (+/-16.2) years (84.2% male). The deep-learning model accurately predicted outcomes of ICU admission/mortality from COVID-19 pneumonia with an AUC of 0.79 (95% CI 0.79-0.96). Compared to traditional risk scoring systems for pneumonia based upon laboratory and clinical parameters, the model matched the EWS and MulBTSA risk scoring systems and outperformed CURB-65.

**Conclusions:** A deep-learning model was able to predict severe patient outcomes (ICU admission and mortality) from COVID-19 on chest radiographs.

**Key Results:** A deep-learning model was able to predict severe patient outcomes (ICU admission and mortality) from COVID-19 from chest radiographs with an AUC of 0.79, which is comparable to traditional risk scoring systems for pneumonia.

**Summary Statement:** This is a chest radiography-based AI model to prognosticate the risk of severe COVID-19 pneumonia outcomes.

## Introduction

The ongoing COVID-19 pandemic continues to be a public health emergency of international concern as declared by the WHO on 30 January 2020 (1). On 23 May 2020 there were 5.21 million cases of infection worldwide and 338,000 deaths had been reported (2). After the easing of extensive shelter-in-place measures, several countries reported a surge in infection rates, raising concerns of a second wave of infections and prompting public health authorities to warn that that the number of cases would spike in the northern hemisphere fall-winter season of 2021 (3).

Morbidity and mortality in COVID-19 infections are dependent on patient factors such as age, burden of chronic illnesses (4), and system factors such as the availability of an adequate healthcare infrastructure for care delivery. Several recent journal articles have described other predictive risk factors such as cancer history, lactate dehydrogenase levels and lymphocyte counts (5,6).

Clinical prognostic scoring tools to aid clinicians in the disposition and severity assessment of COVID-19 have been described (4,7-9). With the use of predictive scores, severity of disease may then be estimated to prompt the clinician to consider the options of ambulatory care, inpatient care, or the use of critical care facilities for severely ill patients. These considerations are becoming more urgent in the current global COVID-19 pandemic due to an unprecedented surge for inpatient care and critical care support. As such, there is an unmet need for unique population-based automated CAP assessment tools that can rapidly stratify disease severity based on a single test. This would enable clinicians to efficiently triage patients focusing on local healthcare resource availability.

One such potential clinical modality is the plain chest radiograph. The efficacy of deep learning-based algorithms based on chest imaging has been demonstrated to be useful for pneumonia and more recently for Covid-19 diagnosis (10-14). Chest radiography has been evaluated for its prognostic utility in predicting COVID-19 outcomes based upon radiologist-generated scoring systems (15). The authors hypothesize that deep-learning-based assessment of chest radiographs can predict severe disease outcomes.

## Methods

### Deep-learning model design

A deep-learning classifier was trained to predict patient prognosis based on chest radiographic images performed on the inpatient admission day.

The deep learning model predicted patient demise and ICU admission during the inpatient episode and combined a pretrained image classification network - Xception with a 30-layer fully-connected network trained on the current data set. Xception is an extension of the inception Architecture which replaces the standard Inception modules with depthwise Separable Convolutions (16).

The use of a predefined model is a transfer learning approach that has the benefit of taking advantage of data from the first setting to extract information that may be useful when learning or even when directly making predictions in the second setting (17). The model was implemented in Keras, version 1.3.0 29 and Scikit-learn, version 0.19.1 and Python, version 3.7 (Python Software Foundation).

### Model development - Training, Validation and Testing

A retrospective study sample of chest radiographs from adult patients admitted to **[blinded for submission] hospital** from 1 January 2019 to 31 December 2019 was used for the development of the predictive model (Figure 1). This retrospective sample utilized chest radiographs from a total of 2,337 inpatient episodes from 1,963 unique adult patients. The target outcome label was defined as a discharge disposition of “deceased” in electronic medical records or ICU admission. A prospective study sample of 70 patients was used to test the model from 1 Jan 2020 to 30 April 2020. The chest radiographs were not selected to exclude other radiographic abnormalities.

**Figure 1:**
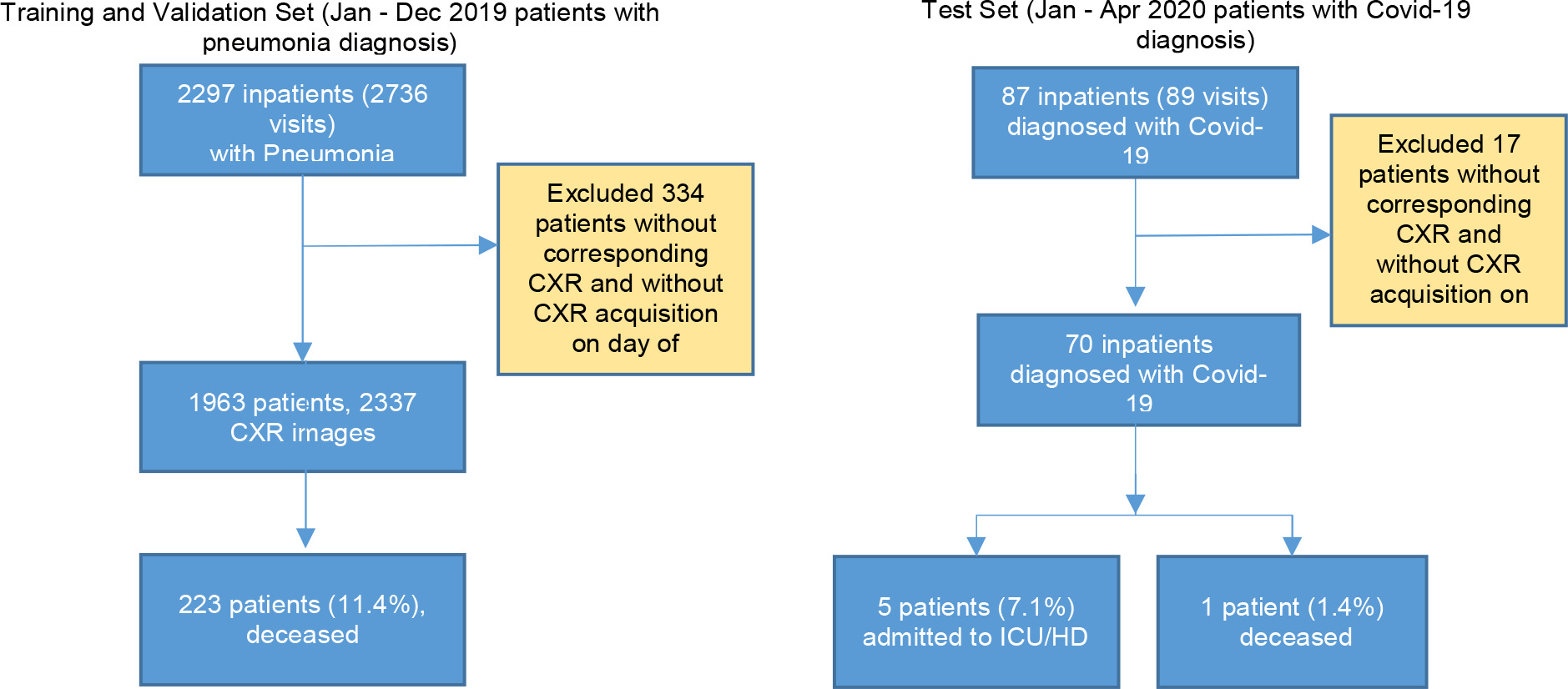
Flowcharts of study sample enrolment. The training and validation sets were sampled retrospectively.

**Figure 2:**
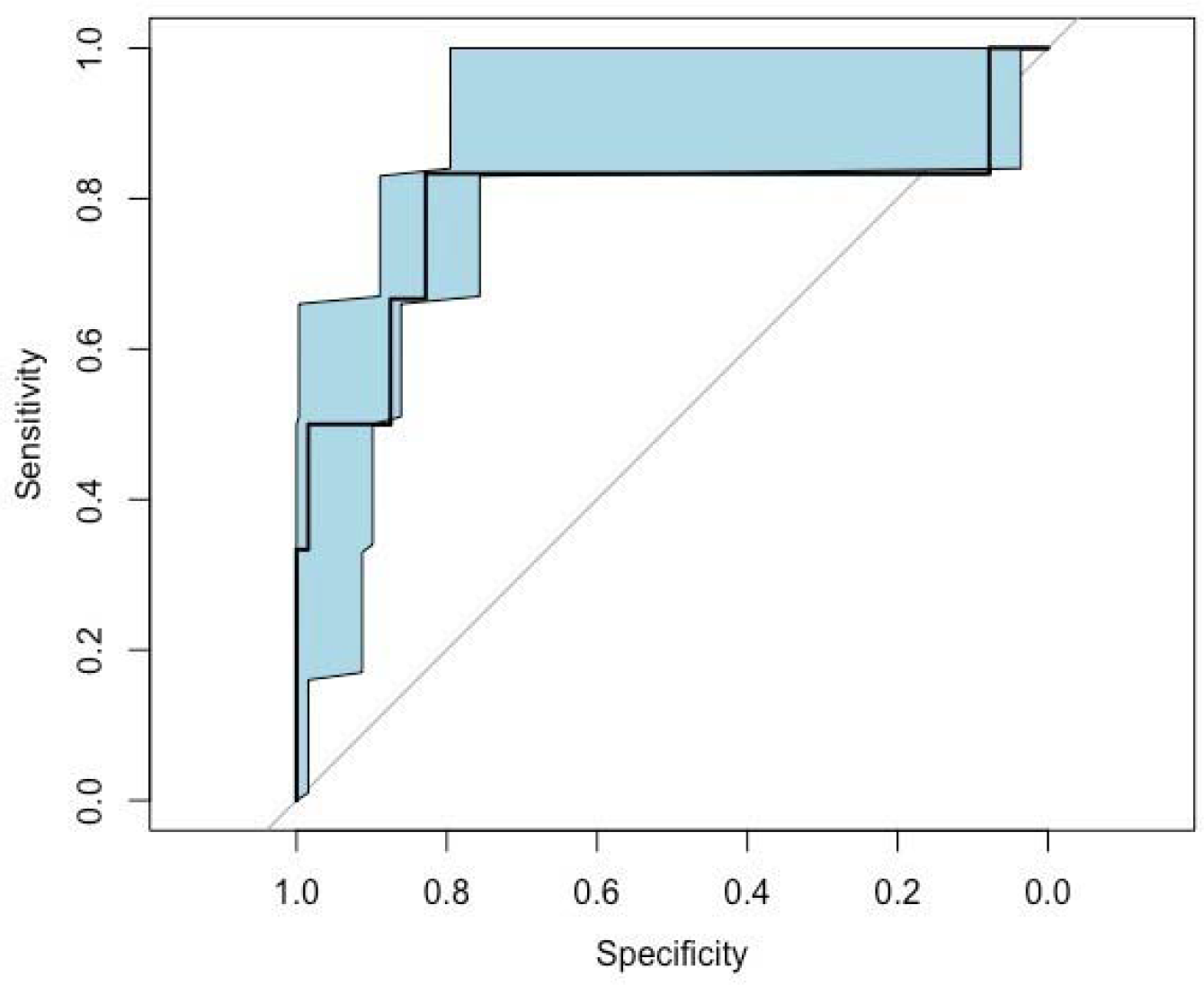
ROC curve for the predictive deep-learning model. 95% confidence intervals are shown as a shaded area for the ROC curve.

Three separate patient sample sets, training, validation and test sets were created (18,19). The training and validation study samples were of patients with an inpatient admission diagnosis of pneumonia as identified by ICD-10 coding and admitted between 1 January 2019 to 31 December 2019. In a situation where there was an overlap between the training and validation set, for example, if chest radiographs belonging to the same patient was found in both the training and validation set, the data was moved from the validation set to the training set.

A split-sample validation was performed. For the training and validation set, we randomly selected 90% [n=2103] of inpatient episodes for model training and held out 10% for validation. The 10% [n= 234] validation set was used for tuning and optimising hypermeters such as dropout rate and learning rate.

Testing of the model was performed with a prospective patient sample of 70 patients with RT-PCR confirmed COVID-19 from 1 Jan 2020 to 30 Apr 2020 [n=70]. The target prediction outcome was defined as mortality and ICU admission.

### Statistical Analysis

Performance of the deep-learning model was assessed by generating receiver operating characteristic (ROC) curves from the model scores, with 95% confidence intervals generated by bootstrapping the observations on the test data. We defined separate operating points for different risk thresholds and computed performance metrics such as sensitivity, specificity, positive predictive value (PPV), negative predictive value (NPV).

## Results

A total of 2,407 inpatient episodes were sampled in the training, validation and testing of the model. The overall prevalence of inpatient episodes associated with in-hospital mortality was 9.3%. The patient characteristics of the training, validation and test sets are summarized in Table 1.

**Table 1:** Patient characteristics of training, validation and test sets. The total number, sex, age, mean length of stay, number of mortalities and number of ICU/HD admissions *ICU, Intensive care unit; HD, High dependency unit; SD, Standard deviation.

| Patient Characteristics | Training & Validation set<br>(Pneumonia diagnosis from<br>Jan- Dec 2019) | Test Set<br>(Covid-19 diagnosis from Jan-Apr 2020) |
| --- | --- | --- |
| Number | 2337 | 70 |
| Age, Mean (SD) [range] | 72 (17.0) [10,100] | 46 (16.2) [23,81] |
| Male (n) / Female (n) | 1311/1026 | 59/11 |
| Mean length of stay (SD)<br>[range] | 5 (6.3) [0,89] | 9 (8.2) [0,35] |
| Number of Mortalities | 223 | 1 |
| Number of ICU/HD | 187 | 5 |

**Table 2:** summary of the predictive model’s performance. AUC = Area under receiver operating characteristic curve; ICU = intensive care unit

|  | AUC (95% CI) |
| --- | --- |
| Model performance for<br>prediction of mortality and ICU<br>admission | 0.794 (0.786 – 0.963) |

The deep-learning system was applied successfully to all 2407 cases. The deep learning model achieved an AUC of 0.79 (95% CI [0.79 –0.96]). The model achieved an accuracy of 93%, specificity of 97% and and NPV of 95% as shown in Table 3.

**Table 3:**
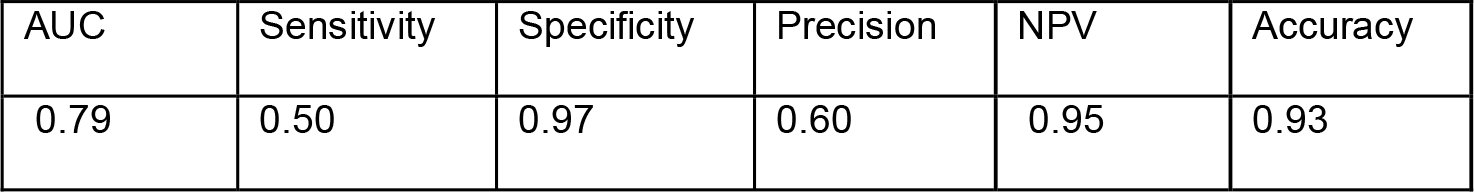
Performance of the predictive deep-learning model on the independent test set. AUC=area under receiver operating characteristic curve, NPV=negative predictive value.

| AUC | Sensitivity | Specificity | Precision | NPV | Accuracy |
| --- | --- | --- | --- | --- | --- |
| 0.79 | 0.50 | 0.97 | 0.60 | 0.95 | 0.93 |

Results of the analysis of accuracy, specificity and NPV are shown in Table 3. We demonstrated performance metrics at different risk thresholds of the model (Table 4). At a risk threshold of 0.5, the specificity was 97%, negative predictive value was 95% and accuracy was 93%. At a risk threshold of 0.6, the specificity was 98%, negative predictive value was 95% and accuracy was 94%.

Figure 3 demonstrates AI generated heatmaps of COVID-19 patients overlaid on chest radiographs, one of a patient who required ICU admission and another of a patient who was not admitted to ICU during the inpatient stay.

**Table 4.**
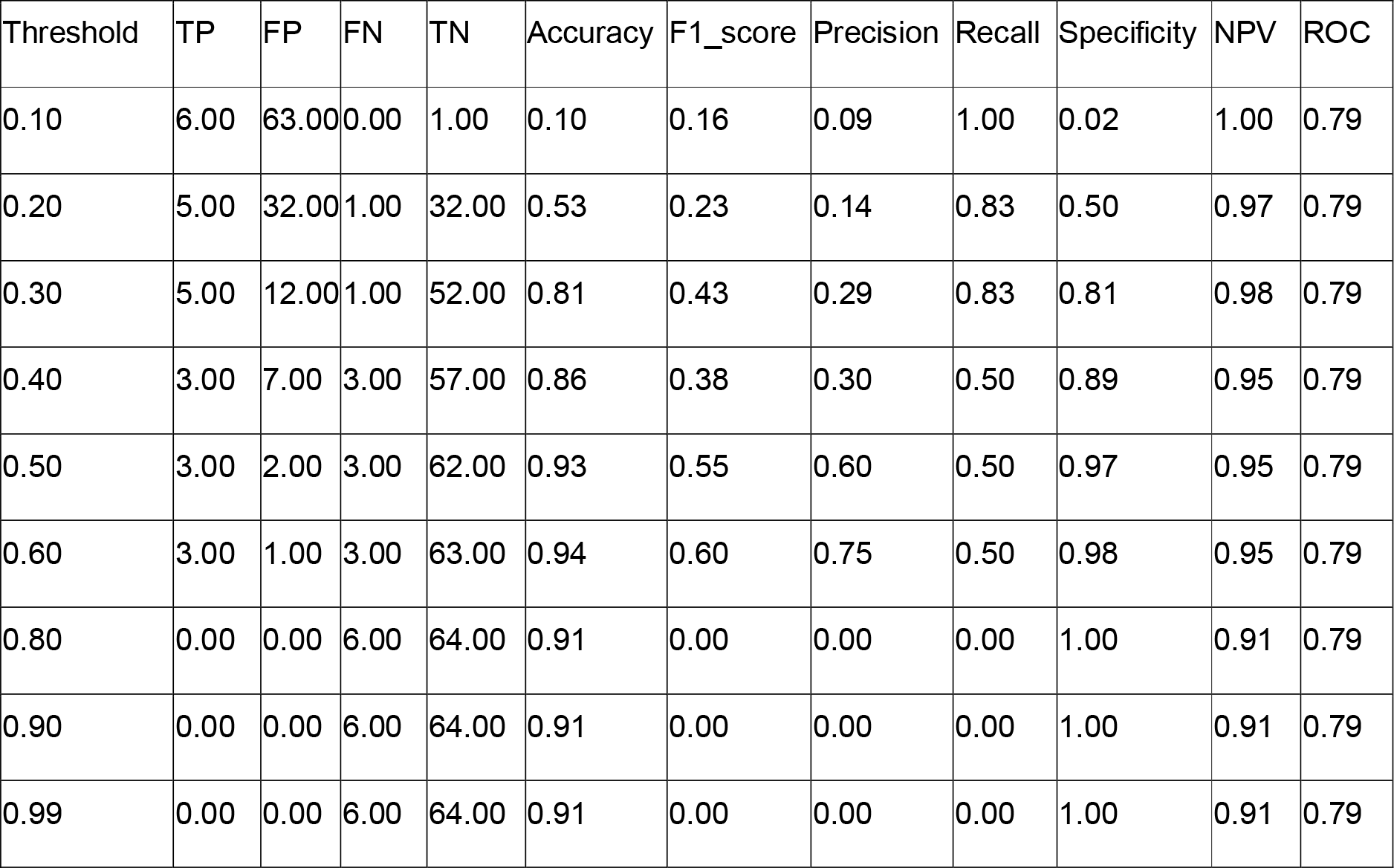
Performance metrics at different risk thresholds of the model. TP=True positive, FP=False positive, FN=False negative, TN=True negative, NPV=Negative predictive value, ROC= receiver operating characteristic

**Figure 3.**
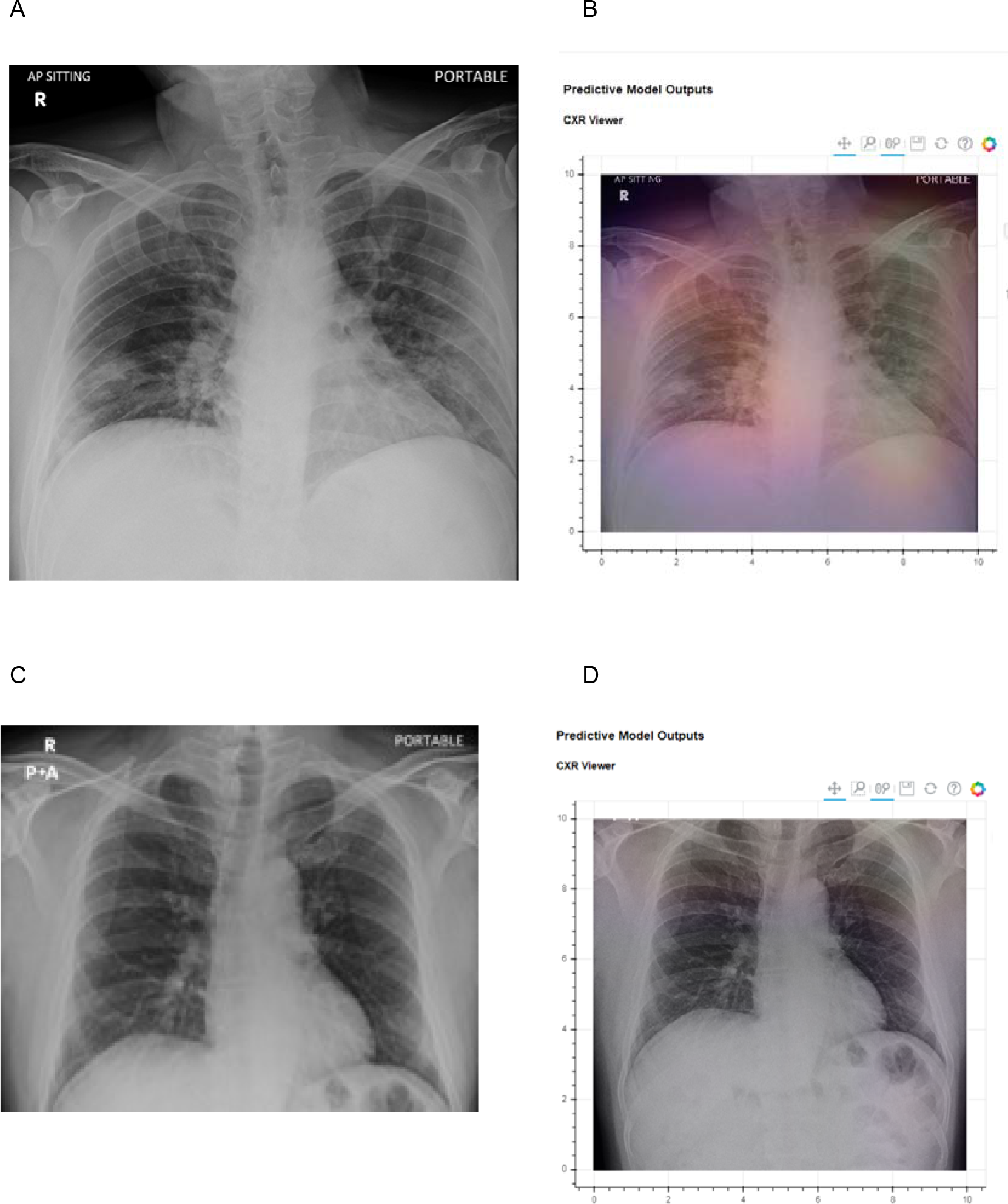
Top row: A 42-year old man with a positive RT-PCR test for SARS-COV2 viral infection. (A) Frontal chest radiograph upon admission (B) The deep-learning model heatmap is overlaid over the image showing pneumonia-related features. The deep-learning model risk score ICU admission for this subject was 80.6%. He was subsequently admitted to ICU for deteriorating oxygen saturation. Bottom Row: A 47-year old man with a positive RT-PCR test for SARS-COV2 viral infection. (C) Frontal chest radiograph upon admission (D) The deep-learning model heatmap is overlaid over the image. The deep-learning model score for this subject was 17.8%. He did not require ICU admission during his admission.

### Generating explainable results of deep-learning interpretation by the utilization of heatmaps

To help the clinician visually interpret how the predictive score was generated by the deep-learning model, we adopted the use of a gradient-weighted class activation map (Grad-CAM) to generate a heatmap to demonstrate the neural-network activated in the forward-pass during inference/prediction. (Figure 3)

## Discussion

In this study, we evaluated the performance of a deep-learning model (Covid-19 AI predictive engine - CAPE) to predict patient outcomes from COVID-19 positive chest radiographs on independent validation and test sets. The results demonstrate that a deep-learning model can be used to predict outcome severity in patients infected with COVID-19 in terms of progression to ICU admission and inpatient mortality. The system’s performance for prediction of COVID-19 outcomes of mortality and ICU admission AUC bootstrapped to our model was 0.79.

Clinical integration of this system would potentially allow improved patient care by providing decision support pertaining to the appropriate disposition of patients to the ambulatory, inpatient or critical care setting at the point of admission. The utilization of a single modality: chest radiography, is quick, relatively inexpensive, and widely available, even in resource-constrained countries.

Notably, the performance achieved by our COVID-19 predictive deep-learning model was comparable to other traditional severe-outcome risk scoring systems developed for viral pneumonia (MuLBSTA) and community acquired pneumonia (CURB-65, EWS): the average AUC of the bootstrapped MuLBSTA model and CURB-65 score were 0.81 and 0.73 respectively (20). Our model outperformed the CURB-65 risk scoring system for ICU admittance, and matched the EWS risk scoring for ICU admittance, with AUC of 0.79 and 0.65 for EWS and CURB-65, respectively (21).

We identified several limitations in our study. Firstly, the training and validation sets were obtained from a single institution, which may not be representative of other institutions. Secondly, the number of RT-PCR confirmed COVID-19 chest radiographs in the test set was relatively small (n=70), relative to the number of non-COVID-19 pneumonia images (n=2736).

To improve the performance of the deep-learning predictive model for COVID-19, a large training dataset of RT-PCR confirmed COVID-19 chest radiographs is needed. The model could also be improved by adopting a multimodal approach by combining radiographic imaging data with clinical and laboratory results for model training.

In conclusion, we evaluated a deep-learning AI model (CAPE: COVID-19 AI Predictive Engine) for the prediction of COVID-19 severe outcomes, namely ICU admission and mortality. The CAPE AI model’s performance was comparable and at times superior to traditional prognostic risk scoring systems for pneumonia. This tool will be made available in the future free-of-charge on the author’s website upon request, to aid in the global health response to COVID-19.

## Data Availability

not applicable

## Abbreviations

Al: Artificial Intelligence
RT-PCR: Reverse Transcription Polymerase Chain Reaction
ROC: Receiver Operating Characteristic
AUC: Area Under Receiver Operating Characteristic Curve
CXR: Chest radiograph

